# The effects of intracranial stimulation on local neurovascular responses in humans

**DOI:** 10.1101/2025.09.09.25335363

**Authors:** Artur Vetkas, Srdjan Sumarac, Samantha Chau, Reese Clinton, Emily Haniff, Alexandre Boutet, Can Sarica, Xilin Liu, Mojgan Hodaie, Taufik A Valiante, Suneil K Kalia, Andres M Lozano, William D Hutchison, Luka Milosevic

**Author notes:** These authors contributed equally to this work (Artur Vetkas, Srdjan Sumarac). **Correspondence:** Luka Milosevic, 399 Bathurst St. 11MP301, Toronto ON, M5T 1S8.

## Abstract

**Background:** Deep brain stimulation (DBS) is used to treat neurological and psychiatric disorders by modulating neuronal circuits. However, other mechanisms of DBS, such as the effects of electrical stimulation on the neurovascular unit remain poorly understood due to limitations of capturing microvascular changes near implanted leads. This motivated us to investigate electrophysiological surrogates of vascular dynamics in response to intraoperative microstimulation.

**Methods:** Microelectrode recordings (n = 193; 108 patients) were obtained during DBS implantation surgery from two electrodes (∼600 µm apart) before and after microstimulation through one electrode. Linear mixed models assessed cardioballistic waveform (CBW) amplitude changes following low-frequency (LFS, 1 Hz) or high-frequency stimulation (HFS, 100 Hz), with comparisons across basal ganglia regions and in white matter. An analytical model interpreted CBW amplitudes as pressure-driven vessel wall expansion, enabling estimation of stimulation-evoked vasodilation and cerebral blood flow.

**Results:** CBW amplitudes increased significantly after HFS at the stimulating electrode, but not at the non-stimulating electrode or after LFS. Significant region-specific effects were observed in the ventral intermediate nucleus (Vim; 107 ± 13%), subthalamic nucleus (STN; 79 ± 7%), and globus pallidus internus (GPi; 78 ± 8%), but not in white matter (50 ± 14%) or substantia nigra (45 ± 8%). Modelling showed that the mean 88% CBW increase across Vim, STN, and GPi corresponds to an acute increase in cerebral blood flow.

**Conclusion:** Intracranial CBW recordings reveal that high-frequency DBS evokes region-specific vascular responses which can be modelled as substantial increases in local blood flow, establishing CBW amplitude as a novel biomarker of subcortical hemodynamics, and a potential therapeutic modality.

## Introduction

Deep brain stimulation (DBS) is a well-established treatment for several neurological disorders, yet the extent to which it modulates cerebrovascular dynamics within subcortical regions remains poorly characterized^1^. While classically viewed as a neuromodulatory intervention targeting neuronal and synaptic activity^2^, DBS is also known to influence cerebral blood flow through both neurovascular coupling (NVC) and possibly direct vascular mechanisms^3,4^. However, its vascular effects in deep brain structures have received limited investigation, largely due to the inaccessibility in measuring the subcortical microvasculature and the limitations of conventional imaging methods^5–7^. As vascular responses underpin the signals captured by modalities like functional magnetic resonance imaging (fMRI) and positron emission tomography (PET)^8,9^, clarifying how DBS engages the subcortical vasculature is essential for understanding both its mechanisms and its broader physiological impact.

Within the neurovascular unit, pial arteries give rise to astrocyte-ensheathed microvessels that interface closely with neurons, forming a structural and functional nexus that coordinates metabolic exchange and vascular tone (Figure 1)^10–12^. This architecture underlies two principal models of NVC: the feedback model, which attributes increased perfusion to rising metabolic demand for ATP and ion homeostasis^10,13^, and the feedforward model, which proposes that neurotransmitter release during neural activation triggers vasodilation through direct or astrocyte-mediated signaling to vascular smooth muscle^3^. While the dominance of either model remains debated, both converge on the principle that neuronal activity drives vascular responses. This forms the basis for investigating how DBS may influence cerebral blood flow through modulation of synaptic pathways. However, considering the intrinsic excitability of astrocytes, pericytes, and smooth muscle cells, DBS may also engage non-neuronal vascular elements directly in altering cerebrovascular dynamics^14^.

**Figure 1.**
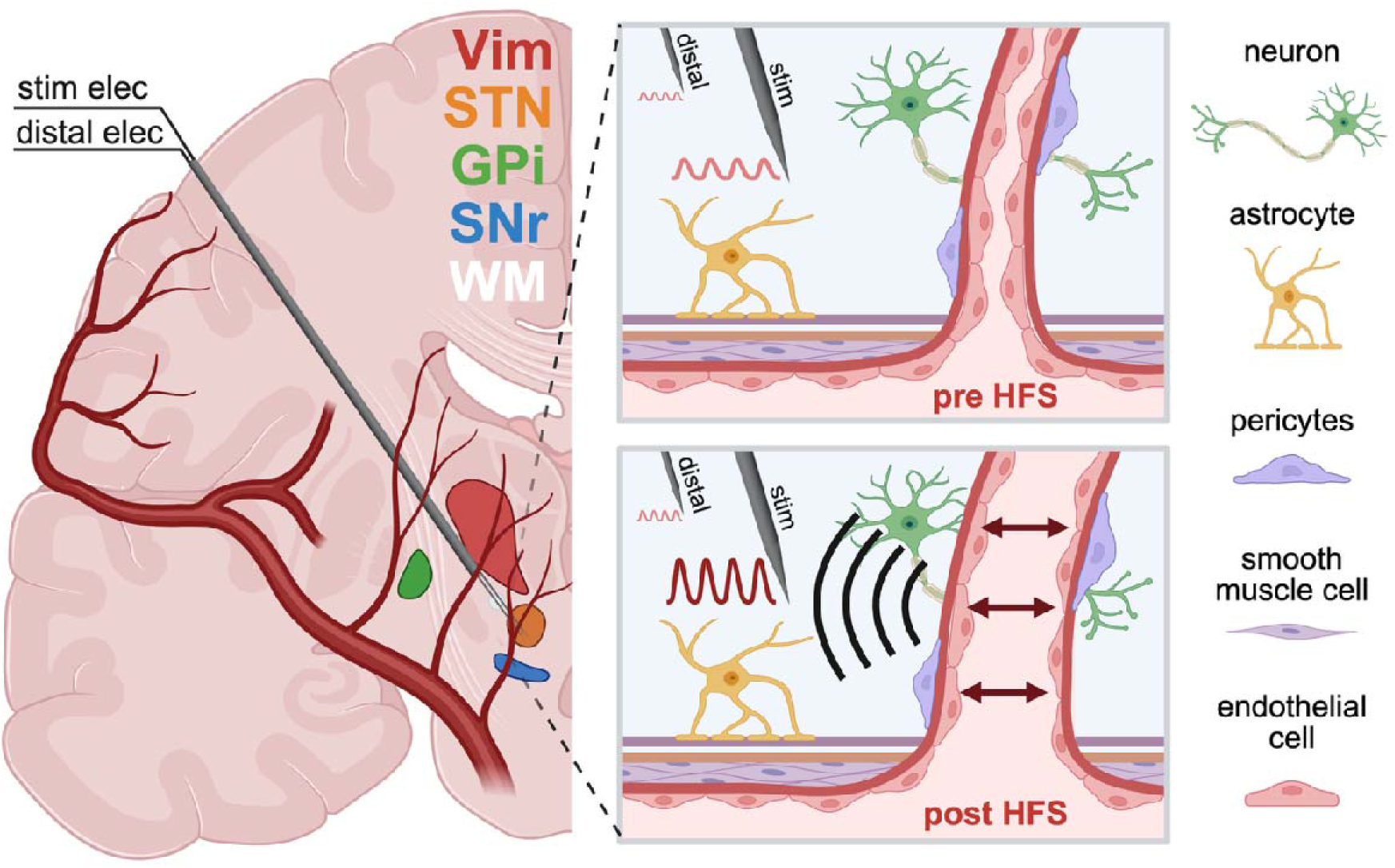
The effect of intracranial stimulation on the NVU. The schematic shows the stimulating and distal recording microelectrodes placed 600 µm apart within the various deep brain regions. Pial arteries transition into arterioles and capillaries, which together with neurons, astrocytes and pericytes compose the NVU to maintain homeostasis. At the level of an arteriole, a microelectrode is shown surrounded by neurons and glial cells, along with smooth muscle cells that regulate the blood vessel diameter and blood flow. The pre- and post-HFS depiction describes the mechanism wherein stimulation-induced vasodilation elevates intraluminal pressure, enhances blood flow, and transmits pulsatility toward the microelectrode. Created with BioRender.com.

Despite growing interest in the effects of DBS on the neurovascular unit and blood-brain barrier (BBB), characterizing how electrical stimulation modulates microvasculature with high spatial and temporal resolution remains challenging. Conventional clinical studies rely on neuroimaging techniques, such as blood oxygenation level-dependent (BOLD) fMRI and PET, to detect stimulation-evoked hemodynamic changes^9,15^; however, these modalities are constrained by limited spatial resolution, limited sensitivity to subcortical vessel dynamics^5^, and vulnerability to stimulation-induced artifacts^7^. Preclinical investigations have provided key mechanistic insights into NVC using optical techniques like two-photon microscopy^16^ and laser Doppler flowmetry^17^. However, these approaches are restricted to superficial cortical layers and require invasive cranial access, limiting their applicability to mostly animal research^18–20^. Notably, a study by Noor and colleagues^21^ demonstrated frequency-dependent cortical vascular responses to thalamic stimulation in rats using intrinsic optical imaging, revealing increased regional blood flow in the motor cortex. While such findings elucidate DBS-induced vascular dynamics at the cortical surface, comparable evidence for deep brain structures in humans remains scarce. This gap highlights the pressing need for alternative approaches capable of resolving rapid microvascular responses within subcortical regions targeted by DBS.

Intracranial recordings offer a unique window into vascular dynamics through physiological signals recorded during stimulation procedures. These recordings often contain cardioballistic signals, rhythmic amplitude modulations phase-locked to the cardiac cycle, arising from vascular pulsations that displace brain tissue relative to the electrode tip^22^, and may also reflect neuronal encoding of cardiac signals in subcortical regions^23^. Although traditionally considered artifacts, these signals may reflect local vascular activity. In this study, we reinterpret the cardioballistic waveform (CBW), a physiological signal arising from cardiac-induced pulsation, as a surrogate biomarker for probing stimulation-evoked vascular responses.

## Methods

### Patients

A total of 193 microelectrode recordings were obtained from 108 patients during deep brain stimulation implantation surgery. High-frequency stimulation (HFS) data were collected from the subthalamic nucleus (STN; n = 32 trajectories, 52 recordings), globus pallidus internus (GPi; n = 27, 49), substantia nigra pars reticulata (SNr; n = 20, 23), ventral intermediate nucleus of the thalamus (Vim; n = 14, 21), and white matter tracts above the subthalamic nucleus (WM; n = 10, 13). Low-frequency stimulation (LFS) recordings were acquired from the STN (n = 12, 13), SNr (n = 13, 17), and GPi (n = 4, 5). The cohort had a mean age of 63.6 ± 9.7 years, with 41.7% female participants. Surgical indications included Parkinson’s disease (n = 94), essential tremor (n = 9), and dystonia (n = 5). All patients provided informed consent prior to study participation. Ethical approval was granted by the Institutional Review Board of the University of Toronto, and the study adhered to principles outlined in the Declaration of Helsinki.

### Microelectrode recordings

Recordings were acquired using two microelectrodes spaced 600 µm apart (impedance 0.05– 0.4 MΩ), with one electrode designated for stimulation and the other, the distal electrode, used exclusively for recording (Figure 1), as previously described^24^. Microstimulation was delivered through the stimulating electrode using either a constant current stimulator (Neuro-Amp1A, Axon Instruments) or the CereStim R96 (Blackrock Microsystems, Salt Lake City, UT, USA). Stimulation parameters included 100 Hz for HFS and 1 Hz for LFS, 100 µA amplitude, 150 µs pulse width, cathode-first polarity, and 2–10 s train durations. The microelectrode recordings (MER) were either amplified using GS3000 Guideline System amplifiers (Axon Instruments) and digitized at a rate of ≥10 kHz with a CED1401 data acquisition system (Cambridge Electronic Design) or acquired using the NeuroPort™ neural signal processor (Blackrock Microsystems, Salt Lake City, UT, USA), sampled at 30 kHz.

### Data processing

To assess vascular reactivity, CBW amplitude changes were measured from MER data. Recordings were segmented into pre- and post-stimulation epochs, each containing at least three identifiable cardioballistic pulses. Post-HFS epochs were initiated ∼2 seconds after stimulation offset to allow recovery from amplifier saturation. Signals were downsampled to 300 Hz after preprocessing with a second-order zero-phase Butterworth anti-aliasing filter (cutoff 150 Hz). Data smoothing employed a Savitzky-Golay filter (window length 31 samples, polynomial order 3). CBW peaks and troughs were identified using a peak detection algorithm (minimum inter-peak interval 0.5 seconds). Median peak-to-trough amplitudes, normalized to pre-HFS local field potential (LFP) root mean square amplitude (bandpass filtered at 10–50 Hz), quantified waveform amplitude changes from pre-to post-stimulation epochs. The percentage change in waveform amplitude from pre-to post-HFS epochs was subsequently computed to quantify the effects of stimulation on vascular reactivity.

### Statistical analysis

Linear mixed models (LMM) were utilized to analyze cardioballistic amplitude changes pre- and post-stimulation. The epoch was included as a fixed effect, while patient ID, recording system (Axon vs Blackrock), stimulation duration, and disease type were random effects. CBW amplitude changes were categorized as increases, decreases, or no change (±10% threshold), presented graphically using a stacked bar chart. Bar graphs depicted amplitude changes across targeted structures, displaying increases or decreases with standard errors.

### Analytical model of CBW as pressure-driven vessel expansion

The cardioballistic waveform (CBW) was modeled as a mechanical signal arising from radial expansion of a nearby vessel in response to pulsatile intraluminal pressure. This expansion displaces surrounding tissue and fluid, generating a detectable signal at the microelectrode. The CBW amplitude was assumed to scale with the vessel wall displacement (*CBW*_amp_ ∝ ΔR). To link this displacement to vascular mechanics, we applied the tube law, which describes how vessel cross-sectional area varies with intraluminal pressure as a function of wall stiffness^25,26^:

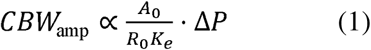

Here, *A*_O_ and *R*_O_ denote the baseline vessel area and radius, *K_e_* is the effective wall stiffness (inversely related to compliance), and Δ*P* is the pulse pressure. To relate CBW to cerebral blood flow (Q), we applied Poiseuille’s law under the assumption of laminar flow and fixed pressure gradient (i.e. constant driving pressure across a short segment). Substituting from Equation (1) and assuming *A*_0_ = πR_0_^2^ we derived:

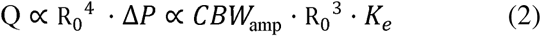

Full derivations for Equations (1) and (2) are provided in the Supplementary Material.

### Data availability

The datasets used and analyzed in this study have been made available in a public repository (see: https://osf.io/mqu5e/)

## Results

### Local and frequency-dependent effects of microstimulation

HFS significantly increased CBW amplitudes at the stimulating electrode (LMM; p < .0001; Figure 2B, part i), whereas amplitudes at the distal electrode did not show significant alterations (LMM; p = .0981; Figure 2B, part ii). LFS also did not result in significant changes in CBW amplitudes (LMM; p = .1723; Figure 2B, part iii).

**Figure 2:**
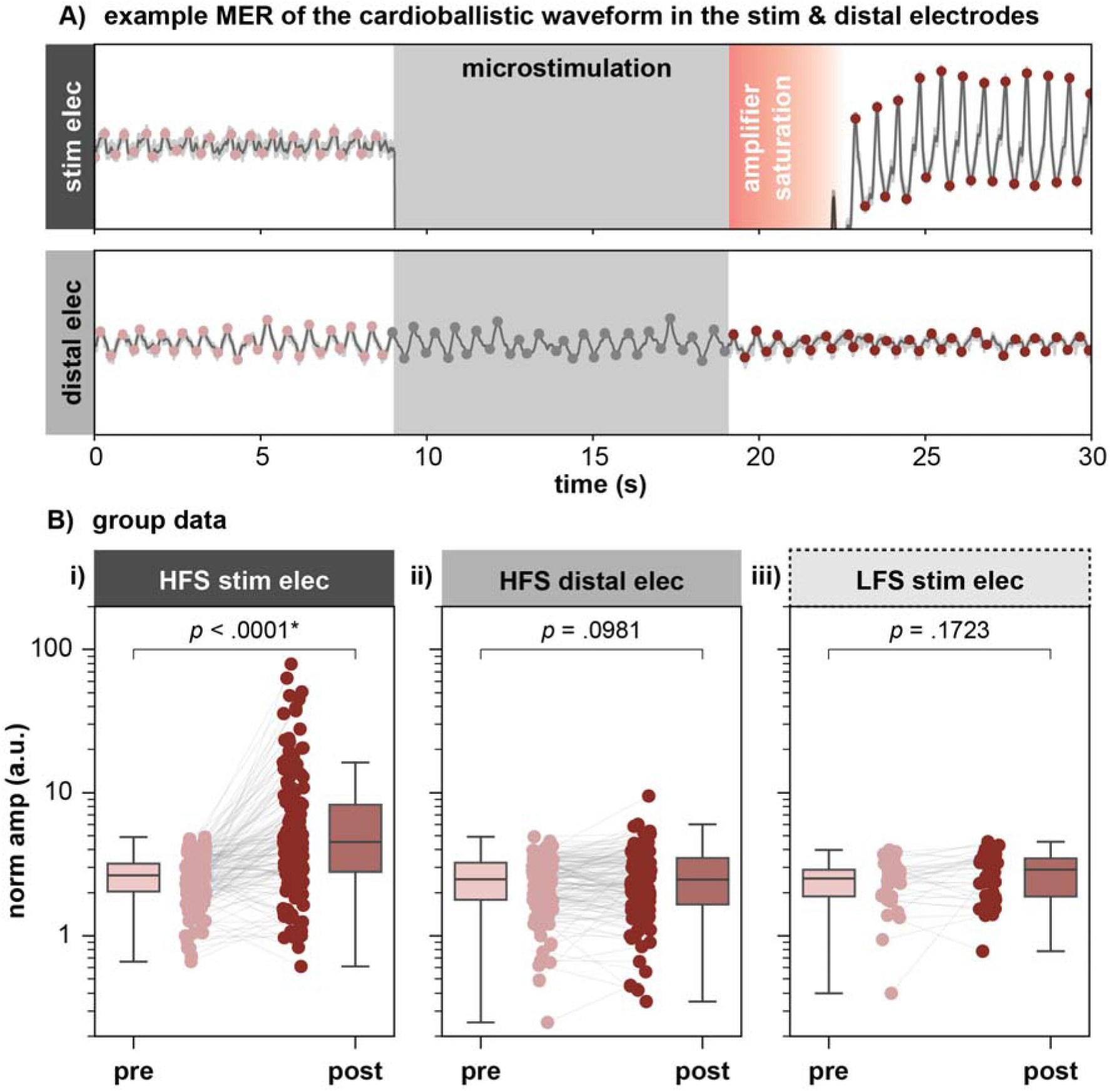
Effects of HFS and LFS on CBW amplitudes. (A) An example trace showing stimulation-induced effects in CBW in the stimulating and distal electrodes. (B) Boxplots comparing CBW amplitudes before and after HFS, demonstrating a statistically significant increase at the stimulating electrode (LMM; p < .0001) but not at the distal electrode (LMM; p = .0981). (C) Boxplots comparing CBW amplitudes before and after LFS reveal no significant effect at the stimulating electrode (LMM; p = .1723).

### Regional variability within the stimulating electrode across gray and white matter

CBW amplitudes exhibited statistically significant increases following HFS in the Vim (p = .0135; Figure 3A, part i), STN (p = .0229; Figure 3A, part ii), and GPi (p = .0002; Figure 3A, part iv). No significant changes were observed in the SNr (p = .0742; Figure 3A, part iii) or white matter (WM) tracts above the STN (p = .1299; Figure 3A, part v). In the Vim, 85.7% of sites showed increased cardioballistic responses, 4.8% decreased, and 9.5% were unchanged; in the STN, 75.0% increased, 17.3% decreased, and 7.7% were unchanged; in the SNr, 73.9% increased, 21.7% decreased, and 4.3% were unchanged; and in the GPi, 71.4% increased, 10.2% decreased, and 18.4% were unchanged. In WM, 53.8% of sites increased, 15.4% decreased, and 30.8% were unchanged (Figure 3B). Mean amplitude increases were 107 ± 13% in Vim, 79 ± 7% in STN, 78 ± 8% in GPi, 45 ± 8% in SNr, and 50 ± 14% in WM (Figure 3C).

**Figure 3:**
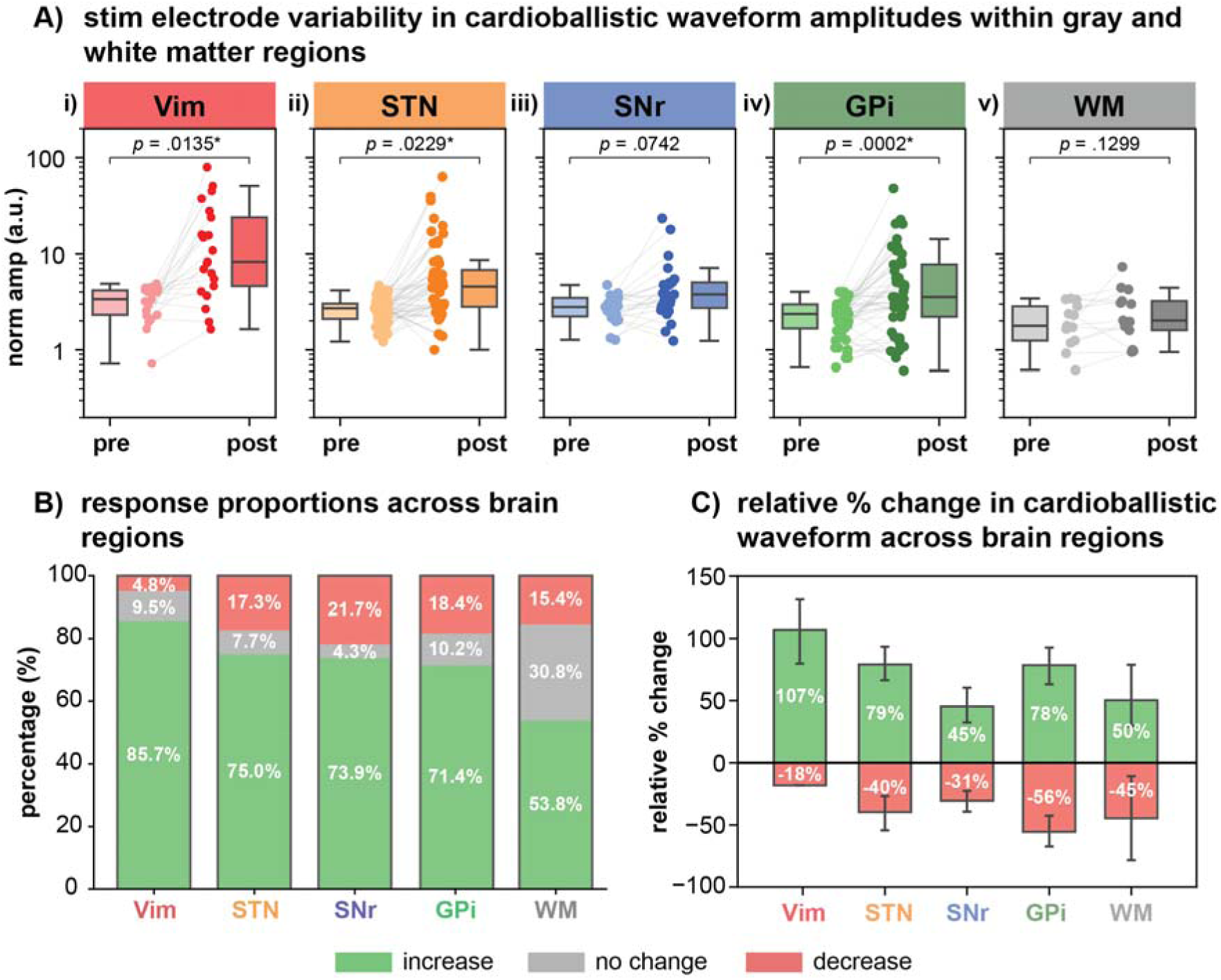
Variation in CBW amplitudes at the stimulating electrode across gray and white matter under HFS. (A) CBW amplitude variability is shown at the stimulating electrode for gray and white matter regions where significant increases are observed in Vim (p = .0135) STN (p = .0229) and GPi (p = .0002) while no significant changes are noted in the SNr (p = .0742) or in the WM region above the STN (p = .1299) (B) A stacked bar chart illustrates the proportion of recordings exhibiting increased (green) decreased (red) or unchanged (gray) amplitudes following HFS (C) A bar graph presents the mean percentage of amplitude increases or decreases after HFS among these structures

**Figure 4:**
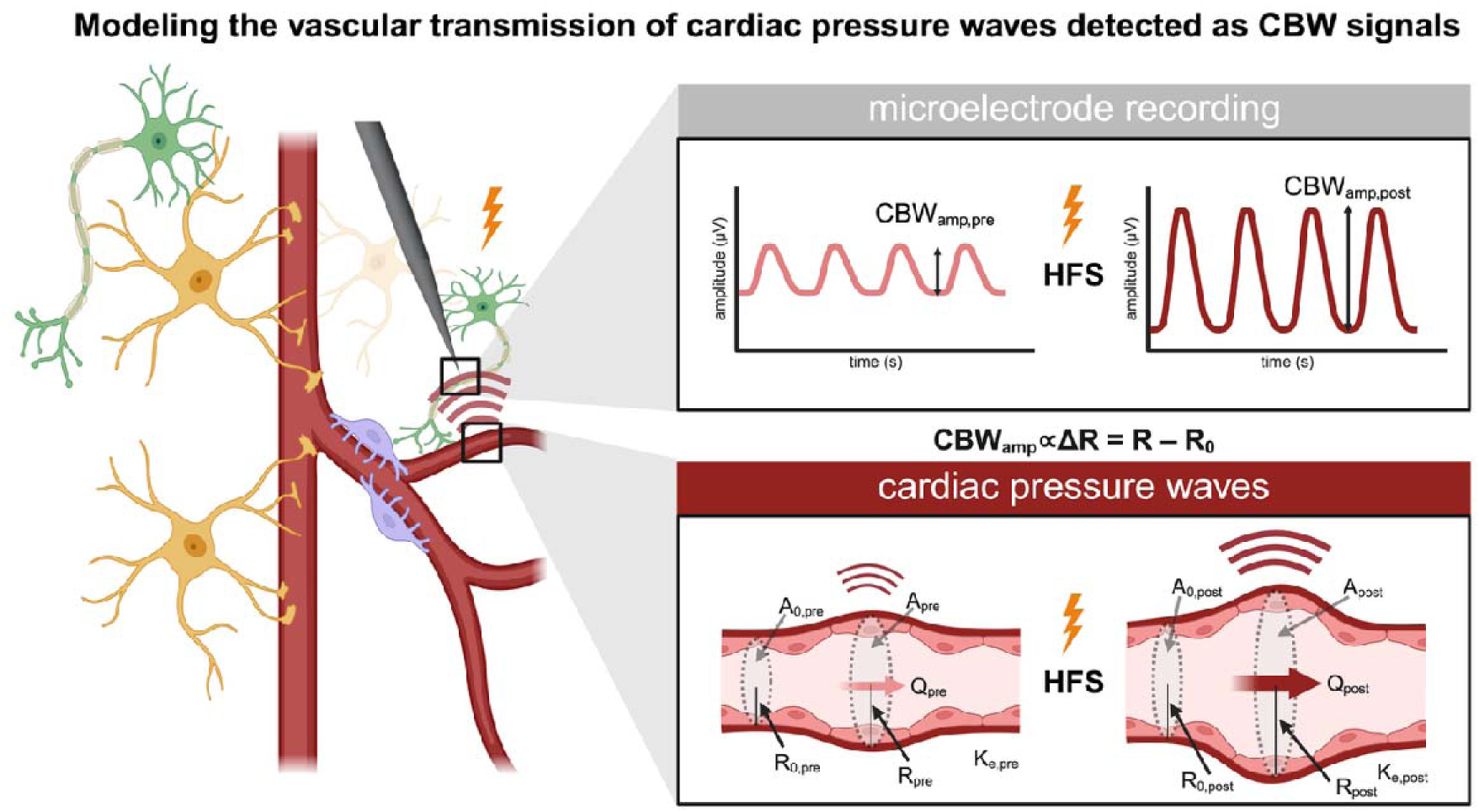
Analytical model linking stimulation-evoked vascular dynamics to CBW signals via pressure-driven wall expansion. Schematic illustrates how cardiac pulsatility induces vessel wall displacement, modeled as a mechanical response to pressure-driven radial expansion. HFS evokes local vasodilation and stiffness reduction, amplifying radial displacement (ΔR = R – R_0_), which manifests as increased CBW amplitude (CBW_amp ∝ ΔR) in microelectrode recordings. The relationship between pressure, vessel mechanics, and CBW signals is derived from the tube law and Poiseuille’s law, establishing a link between stimulation-evoked CBW changes and cerebral blood flow (denoted Q). Based on observed CBW amplitude increases (∼88%), the model estimates a ∼1.7-fold increase in local cerebral blood flow. Created with BioRender.com.

### Analytical estimation of stimulation-evoked hemodynamic changes

To determine whether the observed increases in CBW amplitude within DBS targets reflect physiologically plausible changes in cerebral blood flow, we applied an analytic model derived from vascular mechanics. We assumed that DBS-evoked vasodilation entails a 5% increase in arteriole vessel radius and a 20% reduction in wall stiffness, consistent with documented neurovascular responses to neural activation and modulation^27–29^. Using an average CBW amplitude increase of 88%, calculated across the Vim (107 ± 13%), STN (79 ± 7%), and GPi (78 ± 8%) (Figure 3C), we estimated the corresponding change in local blood flow. Under these physiological assumptions, the CBW increase was consistent with a ∼1.7-fold augmentation in cerebral blood flow across DBS targets. This level of flow enhancement aligns with empirically supported vascular responses observed during neurovascular or pharmacologically induced dilation^30,31^.

## Discussion

This study demonstrates that intracranially recorded CBW, reflecting rhythmic vascular pulsations at the electrode-tissue interface, can be leveraged to probe DBS-evoked vascular dynamics in subcortical regions. High-frequency stimulation elicited significant increases in CBW amplitude at the stimulating electrode, with mean increases of 107% in the Vim, 79% in the STN, and 78% in the GPi, while no significant effects were observed during low-frequency stimulation, or in the SNr, white matter, or at a distal site 600 µm away. These region- and frequency-dependent effects indicate that CBW amplitude is sensitive to localized stimulation-evoked vascular changes. Together, these findings offer a novel electrophysiological approach for investigating neurovascular and potentially non-neuronal mechanisms of DBS within deep brain circuits and motivate further exploration of its utility as a biomarker for subcortical hemodynamics. Based on our modeling, the observed augmentation in CBW amplitude is predicted to correspond to increased local cerebral blood flow, a relationship that merits further evaluation as a potential therapeutic avenue.

### Potential neuronal mechanisms driving regional vascular responses

An enhancement in the CBW observed after microstimulation could be explained either through the effects of NVC, or through direct stimulation of the vessel wall and supporting structures. Our finding that HFS increased CBW amplitudes, whereas LFS did not, may provide insight into mechanisms driving vascular responses in subcortical structures (Figure 2B). HFS is known to modulate neuronal firing rates^32^, oscillatory dynamics^33^, and synaptic transmission by activating axons and promoting neurotransmitter release^34,35^. In contrast, LFS delivers pulses less frequently, leading to a lower cumulative level of stimulation-induced neural activity, which may explain its limited vascular effects^2^. The frequency-specific vascular responses we observed may thus reflect differential engagement of neural circuits that vary in their metabolic demand. Region-specific effects further support this interpretation. HFS within the Vim elicited the strongest response, potentially due to its dense cellular structure and glutamatergic input from cerebellar and cortical afferents^36^, which may recruit thalamocortical circuits and elevate metabolic demand. In the STN and GPi, prior work has shown that HFS sustains GABAergic input from the GPe^37,38^, thereby maintaining neurotransmitter release and activating the broader basal ganglia network, which could similarly heighten local energy demands. In contrast, the SNr shows synaptic fatigue and breakthrough spiking during HFS^35,39^, which may decouple it from upstream activity and reduce local metabolic burden. While both the GPi and SNr receive major input from the striatum and GPe, the relative contribution may differ, with potentially greater GPe influence in the GPi than in the SNr^2^. White matter showed no effect, which is consistent with its limited synaptic activity and lower metabolic and vascular support, with microvascular density reported to be 20–49% lower than in cortical gray matter in aged human brains^40^. Additionally, white matter exhibits markedly lower capillary and arteriole density, with capillary length density at 160 mm/mm³ compared to 256 mm/mm³ in cortex and 328 mm/mm³ in subcortex, reflecting its sparser microvascular architecture^41^. Together, these findings suggest that stimulation-induced vascular responses reflect the degree to which DBS engages metabolically active circuits, linking regional neurophysiology to local hemodynamics.

### Potential non-neuronal contributions to DBS-evoked vascular changes

While neuronal signaling may initiate neurovascular responses by increasing metabolic demand, our findings raise the possibility that DBS-induced vascular effects also involve direct modulation of non-neuronal vascular elements. This interpretation is supported by the focal, frequency-dependent CBW increases observed at the stimulating electrode but not at a nearby site 600 µm away (Figure 2B), suggesting localized engagement of vascular structures beyond metabolic neuronal effects. Notably, although our previous studies have demonstrated robust neuronal responses at this distal site (including neuronal inhibition and evoked potentials^34,37^) the absence of a CBW effect here suggests that the threshold for direct vascular activation is likely higher than that required to engage axons or terminals. Several candidate cell types within the neurovascular unit may contribute to these effects. A recent study by Mishra and colleagues directly investigated the stimulation-evoked capillary dilation using two-photon microscopy in rat brain slices. 100 Hz trains of 200 ms were applied using a CSF-filled patch pipette electrode, and electrodes placed 400-500 µm from the imaged vessel for 5 or 60 seconds, with a set up and stimulation settings similar to the microstimulation used herein. The electrical stimulation led to capillary dilation, which was found to be mediated by astrocyte Ca2+ signaling and dependent on postsynaptic activity. Blocking ionotropic glutamate receptors (AMPA/KA) inhibited the dilation, while NMDA receptor antagonists had no effect^16^. Pericytes, which possess excitable membrane properties, may also respond directly to electrical stimulation, as demonstrated in retinal tissue where stimulation evoked contraction independent of synaptic input^42^. Additionally, DBS may influence vascular tone by depolarizing smooth muscle cells lining arterioles, consistent with reports of stimulation-induced increases in microvascular perfusion via calcium-driven mechanisms^43,44^. Although less well established in subcortical regions, endothelial cells may contribute through nitric oxide-mediated vasodilation, given that peripheral electrical stimulation has been shown to enhance flow in an endothelium-dependent manner^45^. Finally, the graded vascular responses observed across basal ganglia targets may reflect region-specific differences in perivascular architecture, including capillary density, astrocytic coverage, and cellular heterogeneity within the neurovascular unit^46^.

### Therapeutic potential and translational implications

These findings suggest that targeted neuromodulation strategies may enhance neurovascular function by modulating neural circuits and potentially vascular elements, offering therapeutic potential in conditions such as ischemic stroke, cerebral vasospasm, and neurodegenerative disease^47–49^. Altered neurovascular coupling and pathologies of BBB have been implicated in disorders like Parkinson’s disease, Alzheimer’s disease, and depression, where DBS may improve symptoms by restoring network activity and metabolic demand^50^. Moreover, electrical stimulation has been shown to modulate BBB properties, including tight junction integrity and transcytosis, which may have implications for neuroinflammatory modulation and drug delivery^51^. BBB polarization and change in electric field density surrounding microvasculature, described by finite-element models simulating DBS-level fields, suggests that DBS could directly drive electroosmotic transport across endothelial walls, boosting metabolic supply and interstitial-fluid clearance beyond standard NVC mechanisms^52^. These insights could inform the design of next-generation minimally invasive brain-computer interfaces, such as endovascular electrodes capable of targeting deep structures or vascular segments without highly invasive surgery^3,53–56^. Finally, by restoring vascular and BBB reactivity, stimulation strategies may mitigate perfusion deficits in critical conditions such as subarachnoid hemorrhage, traumatic brain injury, or the post-stroke penumbra^57,58^.

### Limitations and future directions

Several limitations should be considered when interpreting our findings. First, the CBW remains an indirect proxy for local vascular dynamics and may not fully capture the magnitude or nature of DBS-induced changes in vessel diameter or flow. Future work employing direct hemodynamic modalities, such as transcranial Doppler ultrasound or optical imaging, will be essential to validate and refine this surrogate. Second, our study did not monitor systemic physiological variables such as cardiac output or blood pressure, which could potentially influence CBW amplitudes. Incorporating real-time cardiovascular monitoring in future protocols would help disentangle central and peripheral contributions to the observed effects. Methodological differences between recording systems also warrant consideration. The Axon system applied a built-in, non-disableable 10 Hz one-pole high-pass filter (with shallow roll-off and weak attenuation below cutoff), which likely affects low-frequency components of the cardioballistic signal. In contrast, the Blackrock system recorded unfiltered raw data. Despite this discrepancy, subgroup analyses confirmed consistent proximal effects across both systems (Supplementary Figure 1). Additionally, the relatively short stimulation epochs precluded assessment of long-term CBW dynamics, which could yield further insight into temporal patterns of neurovascular modulation. Lastly, variability in vascular reactivity across subcortical regions may reflect underlying anatomical or pathological differences in neurovascular unit composition. Future studies incorporating longer recordings, broader stimulation parameters, and multimodal validation are needed to more comprehensively characterize the mechanisms underlying DBS-induced vascular responses. Addressing these limitations may help establish electrophysiological surrogates as viable tools for guiding therapeutic strategies that enhance cerebral perfusion in neurological disease^43^.

### Conclusion

This study identifies stimulation frequency- and site-specific increases in CBW amplitude as a novel electrophysiological marker of subcortical vascular reactivity during DBS. HFS, but not LFS, elicited localized effects at the site of stimulation within gray matter, absent in white matter and in distal recording locations. Although CBW remains an indirect measure, the findings suggest that DBS may modulate the neurovascular unit through neurovascular coupling and/or direct engagement of the vascular units. Analytical modeling supports a mechanistic interpretation in which CBW amplitude scales with pressure-driven vessel wall expansion, consistent with DBS-evoked vasodilation and physiologically plausible increases in blood flow. These insights highlight previously underappreciated therapeutic dimensions of DBS and vascular stimulation and motivate further validation using direct hemodynamic techniques.

## Acknowledgments

This project has been made possible with the financial support through the Natural Sciences and Engineering Council (NSERC) RGPIN-2022-05181 (L.M.); Canadian Institute for Health Research (CIHR) PJT 191880 (L.M., S.K.K.).

