## Supplementary for "The effects of intracranial stimulation on local neurovascular responses in humans"

**Supplementary Table 1**

| **disease** | **patient** | **mean age (yrs)** | **% female** | **STN** | **GPi** | **Vim** |
| --- | --- | --- | --- | --- | --- | --- |
| Parkinson’s disease | 94 | 62.4 | 33.23% (35/94) | 65 | 27 | 2 |
| essential tremor | 9 | 75.4 | 55.56% (5/9) | 0 | 0 | 9 |
| dystonia | 5 | 66.1 | 80% (4/5) | 0 | 2 | 3 |

| **patient ID** | **sex** | **DBS target** | **disease** |
| --- | --- | --- | --- |
| 001 | female | GPi | Parkinson's disease |
| 002 | male | STN | Parkinson's disease |
| 003 | male | STN | Parkinson's disease |
| 004 | female | STN | Parkinson's disease |
| 005 | female | STN | Parkinson's disease |
| 006 | male | STN | Parkinson's disease |
| 007 | male | STN | Parkinson's disease |
| 008 | male | GPi | Parkinson's disease |
| 009 | male | GPi | Parkinson's disease |
| 010 | male | STN | Parkinson's disease |
| 011 | male | GPi | Parkinson's disease |
| 012 | female | GPi | Parkinson's disease |
| 013 | male | STN | Parkinson's disease |
| 014 | male | STN | Parkinson's disease |
| 015 | female | GPi | Parkinson's disease |
| 016 | male | STN | Parkinson's disease |
| 017 | male | STN | Parkinson's disease |
| 018 | male | STN | Parkinson's disease |
| 019 | female | STN | Parkinson's disease |
| 020 | female | GPi | Parkinson's disease |
| 021 | female | STN | Parkinson's disease |
| 022 | male | STN | Parkinson's disease |
| 023 | male | STN | Parkinson's disease |
| 024 | female | STN | Parkinson's disease |
| 025 | female | GPi | Parkinson's disease |
| 026 | male | STN | Parkinson's disease |
| 027 | male | STN | Parkinson's disease |
| 028 | female | STN | Parkinson's disease |
| 029 | male | Vim | essential tremor |
| 030 | female | Vim | essential tremor |
| 031 | male | Vim | essential tremor |
| 032 | female | STN | Parkinson's disease |
| 033 | male | STN | Parkinson's disease |
| 034 | male | Vim | Parkinson's disease |
| 035 | male | GPi | Parkinson's disease |
| 036 | female | Vim | dystonia |
| 037 | female | Vim | essential tremor |
| 038 | female | Vim | essential tremor |
| 039 | female | Vim | essential tremor |
| 040 | female | Vim | dystonia |
| 041 | male | Vim | Parkinson's disease |
| 042 | female | Vim | essential tremor |
| 043 | female | Vim | essential tremor |
| 044 | female | GPi | Parkinson's disease |
| 045 | female | GPi | Parkinson's disease |
| 046 | female | Vim | dystonia |
| 047 | male | GPi | Parkinson's disease |
| 048 | male | GPi | Parkinson's disease |
| 049 | male | GPi | Parkinson's disease |
| 050 | male | GPi | Parkinson's disease |
| 051 | male | STN | Parkinson's disease |
| 052 | female | STN | Parkinson's disease |
| 053 | female | STN | Parkinson's disease |
| 054 | male | STN | Parkinson's disease |
| 055 | female | GPi | Parkinson's disease |
| 056 | male | STN | Parkinson's disease |
| 057 | male | STN | Parkinson's disease |
| 058 | female | GPi | Parkinson's disease |
| 059 | male | GPi | Parkinson's disease |
| 060 | female | GPi | Parkinson's disease |
| 061 | male | GPi | Parkinson's disease |
| 062 | male | GPi | Parkinson's disease |
| 063 | male | STN | Parkinson's disease |
| 064 | female | GPi | Parkinson's disease |
| 065 | female | GPi | dystonia |
| 066 | male | GPi | Parkinson's disease |
| 067 | female | STN | Parkinson's disease |
| 068 | male | GPi | Parkinson's disease |
| 069 | female | STN | Parkinson's disease |
| 070 | male | STN | Parkinson's disease |
| 071 | female | STN | Parkinson's disease |
| 072 | male | STN | Parkinson's disease |
| 073 | male | GPi | Parkinson's disease |
| 074 | female | STN | Parkinson's disease |
| 075 | female | STN | Parkinson's disease |
| 076 | male | STN | Parkinson's disease |
| 077 | female | STN | Parkinson's disease |
| 078 | male | GPi | dystonia |
| 079 | female | STN | Parkinson's disease |
| 080 | male | STN | Parkinson's disease |
| 081 | female | STN | Parkinson's disease |
| 082 | male | STN | Parkinson's disease |
| 083 | male | STN | Parkinson's disease |
| 084 | male | STN | Parkinson's disease |
| 085 | male | STN | Parkinson's disease |
| 086 | female | STN | Parkinson's disease |
| 087 | male | STN | Parkinson's disease |
| 088 | male | STN | Parkinson's disease |
| 089 | male | STN | Parkinson's disease |
| 090 | male | STN | Parkinson's disease |
| 091 | male | Vim | essential tremor |
| 092 | female | STN | Parkinson's disease |
| 093 | female | STN | Parkinson's disease |
| 094 | male | STN | Parkinson's disease |
| 095 | male | STN | Parkinson's disease |
| 096 | female | GPi | Parkinson's disease |
| 097 | male | STN | Parkinson's disease |
| 098 | male | STN | Parkinson's disease |
| 099 | male | STN | Parkinson's disease |
| 100 | female | STN | Parkinson's disease |
| 101 | male | STN | Parkinson's disease |
| 102 | female | STN | Parkinson's disease |
| 103 | female | GPi | Parkinson's disease |
| 104 | male | STN | Parkinson's disease |
| 105 | male | STN | Parkinson's disease |
| 106 | male | STN | Parkinson's disease |
| 107 | male | STN | Parkinson's disease |
| 108 | male | STN | Parkinson's disease |

**Deriving the relationship between CBW amplitude and pressure-induced vessel expansion**

The cardioballistic waveform (CBW) was modeled as a mechanical response to pulsatile intraluminal pressure $P$, which induces transient radial expansion of a blood vessel. The signal amplitude, denoted ${CBW}_{\text{amp}}$, was assumed to scale with the maximum radial displacement of the vessel wall during each cardiac cycle. To relate the amplitude to vascular properties and pressure dynamics, we began with the tube law, which models the pressure-area relationship for a thin-walled, elastic vessel. In its linearized form (as applied in Zhao et al. and Raines et al.):

$\Delta A\propto\frac{A_{0}}{K_{e}}\cdot\Delta P$	(S1)

Where:

- $\Delta A=A-A_{0}$: the pressure-induced change in cross-sectional area,
- $A_{0}:$the baseline cross-sectional area at diastolic pressure,
- $\Delta P=P_{systolic}-P_{diastolic}$: the pulse pressure,
- $K_{e}$: the effective wall stiffness effective.

Assuming circular geometry, the area is related to the radius by:

$A=\pi R^{2}\Rightarrow$ $\Delta A=\pi\left( R^{2}-{R_{0}}^{2} \right)$	(S2)

Using the identity $R^{2}-R_{0}^{2}=\left( R-R_{0} \right)\left( R+R_{0} \right)=\Delta R\left( R+R_{0} \right)$, we obtain:

$\Delta A=\pi\cdot\Delta R\cdot\left( R^{2}+{R_{0}}^{2} \right)$	(S3)

Assuming small deformations, where $\Delta R\ll R_{0}$, we approximate $R+R_{0}\approx2R_{0}$

$\Delta A\approx2\pi R_{0}\cdot\Delta R$	(S4)

Solving for $\Delta R$:

$\Delta R\approx\frac{\Delta A}{2\pi R_{0}}$	(S5)

Substitute Equation (S1) into (S5):

$\Delta R\approx\frac{1}{2\pi R_{0}}\cdot\frac{A_{0}}{K_{e}}\cdot\Delta P$	(S6)

Since the CBW amplitude is assumed proportional to the radial displacement $\Delta R$, we arrive at:

${CBW}_{\text{amp}}\propto\frac{A_{0}}{R_{0}K_{e}}\cdot\Delta P$	(1)

**Deriving the relationship between CBW amplitude and cerebral blood flow**

To connect the CBW amplitude to cerebral blood flow, we applied Poiseuille’s law for steady, laminar flow through a cylindrical vessel:

$Q=\frac{{{\pi R}_{0}}^{4}}{8\mu L}\cdot\Delta P$	(S7)

Where:

- $Q$: volumetric blood flow,
- $\mu$: dynamic viscosity,
- $L$: length of the vessel segment,
- $\Delta P$: pressure drop across the segment.

Omitting constant terms and assuming constant viscosity and vessel length, this simplifies to:

$Q{{\propto R}_{0}}^{4}\cdot\Delta P$	(S8)

*NOTE: Although Poiseuille’s law defines flow in terms of the mean pressure drop across a vessel segment, and the tube law describes vessel expansion in terms of pulse pressure, these two quantities are functionally related in the context of pulsatile flow. The same intraluminal pressure wave that drives vessel wall motion also produces a pressure gradient that contributes to bulk forward flow. Therefore, we approximate the pressure drop driving flow as being proportional to the pulse pressure that drives radial expansion. This assumption enables us to substitute between frameworks and express blood flow in terms of CBW amplitude.*

Now, solving Equation (1) for $\Delta P$:

${\Delta P\propto CBW}_{\text{amp}}\cdot\frac{R_{0}K_{e}}{A_{0}}$	(S9)

Substitute (S9) into (S8):

${Q{{\propto R}_{0}}^{4}\cdot(CBW}_{\text{amp}}\cdot\frac{R_{0}K_{e}}{A_{0}})={CBW}_{\text{amp}}\cdot\frac{{R_{0}}^{5}K_{e}}{A_{0}}$	(S10)

Assuming circular geometry $A_{0}=\pi{R_{0}}^{2}$ , this further simplifies:

$\text{Q}\propto{CBW}_{\text{amp}}\cdot R_{0}^{3}\cdot K_{e}$	(2)

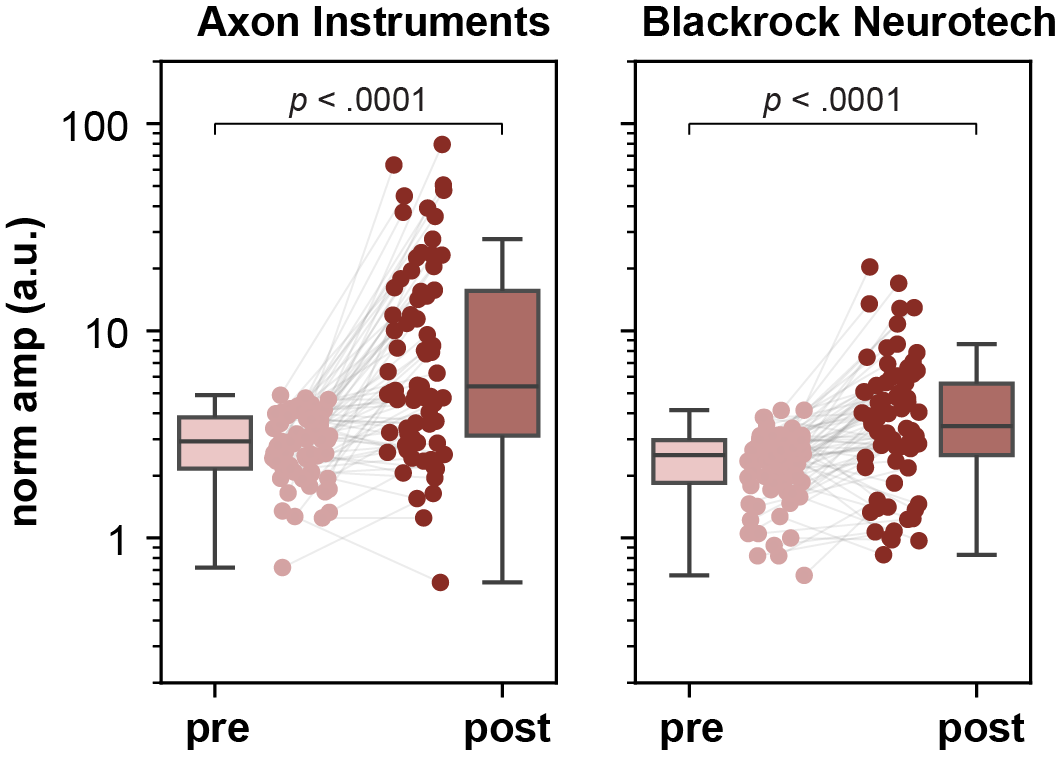

**Supplementary Figure 1: Comparison of cardioballistic waveform amplitudes (pre- and post-HFS in gray matter structures) recorded using the Axon and Blackrock systems.** Significant effects of HFS at the stimulation electrode were observed with both systems (LMM; p < 0.0001 for both).

*Note: The Axon system employed a built-in, non-disableable 10 Hz one-pole high-pass filter, while the Blackrock system recorded unfiltered raw data. Despite the Axon filter's attenuation of low-frequency components, its shallow roll-off partially preserved these frequencies.*
